# Geographic accessibility to public healthcare facilities and spatial clustering during the wet and dry seasons in Côte d’Ivoire

**DOI:** 10.1101/2023.11.21.23298865

**Authors:** Gaëlle T. Sehi, Clarisse A. Houngbedji, Daniel M. Parker, Peter M. Macharia

**Affiliations:** Population Health and Disease Prevention, Public Health, University of California, Irvine, United States; Centre d’Entomologie Médical et Vétérinaire (CEMV), Université Alassane Ouattara, Bouaké, Côte d’Ivoire; Centre Suisse de Recherches Scientifiques en Côte d’Ivoire, Abidjan, 01 BP 1003 Abidjan 01, Côte d’Ivoire; Department of Public Health, Institute of Tropical Medicine, Antwerpen, Belgium; Centre for Health Informatics, Computing and Statistics, Lancaster Medical School, Faculty of Health and Medicine, Lancaster University, Lancaster, United Kingdom; Population & Health Impact surveillance Group, Kenya Medical Research Institute-Wellcome Trust Research Programme, Nairobi, Kenya

## Abstract

**Introduction:** Geographic accessibility to healthcare is a key determinant of health outcomes. Yet, globally, over 4.5 billion people lack access to healthcare, majority of which are located in low and middle-income countries. While metrics of spatial access should consider variation in seasonality, studies in Côte d’Ivoire have overlooked seasonality impacts and how such metrics are spatially clustered. Here, we assessed geographical accessibility to public health care in Côte d’Ivoire across wet and dry seasons while assessing clustering and identifying marginalized populations.

**Methods:** We assembled spatial data on the location of public healthcare facilities, factors that affect travel, population distribution, and healthcare-seeking behaviour. Using these datasets, plausible travel scenarios reflecting seasonality were combined within a cost-distance algorithm in AccessMod (version 5) to model travel time to the nearest facility at 1km resolution for dry and wet seasons. We determined the proportion of the population within 2 hours’ travel time at the district level. We assessed marginalization (≥4 hours from the nearest facility) and spatial autocorrelation using Moran’s I indices.

**Results:** At the national level, mean travel time to the nearest public healthcare facility was 1.8h during the dry season and 3.4h during the wet season. The travel times were heterogeneous and varied between 23min – 6h and 38min - 10h during both seasons. About 73% (7 to 98%) of the population was within 2h of the nearest public healthcare facility in the dry season and 55% (2 – 97%) during the wet season at the district level. Poor access clusters were in the north and better access clusters in the south-central area of the country during both seasons.

**Conclusion:** Healthcare access inequalities in Côte d’Ivoire persist, with inadequate access clusters in the north and better access in the south-central region. There is a need for seasonal-based targeted interventions to improve access.

**Key Summary:** *What is already known on this topic:*

- Adequate geographic access to healthcare is crucial for timely diagnosis and life-saving interventions, especially in low- and middle-income countries where individuals face physical barriers to accessing essential and routine healthcare.
- Healthcare access in Côte d’Ivoire has been examined without considering weather seasonality or spatial clustering, yet these play a critical in healthcare accessibility.

*What this study adds:*

- In the dry season, the mean travel time to the nearest healthcare facility at the national level is approximately 1.8 hours, whereas in the wet season, it increases to 3.4 hours.
- Over 80% of the population can reach a healthcare facility within 2 hours during the dry season, but only 66% can do so during the wet season.

*How this study might affect research, practice or policy.:*

- Policymakers can use this study to develop policies that address seasonal variations in accessibility, ensuring that healthcare services remain accessible even during adverse weather conditions.

## Introduction

Access to healthcare facilities is a strong predictor of health utilization, with implications for both individual and population-level health outcomes. In 2021, approximately 4.5 billion people did not have adequate access to essential health services (World Health Organization [WHO] & Bank, 2023). Healthcare access can be grouped into various taxonomies that include geographic versus social accessibility, or potential versus realized accessibility (Guagliardo, 2004; Penchansky & Thomas, 1981; Shah et al., 2016).

Geographical accessibility refers to the relative ease or difficulty of reaching healthcare services based on the physical location of the individual needing care, the physical location of a healthcare facility or provision point, and the availability of transportation (including transportation costs). Geographic accessibility is directly related to healthcare utilization (Agbenyo et al., 2017; Carrasco-Escobar et al., 2020; Kanuganti et al., 2015) which in turn affects health outcomes such as immunization rates, coverage of interventions, and child mortality (Blanford et al., 2012; Makanga et al., 2017; Tanou et al., 2021). Additionally, access to timely diagnosis and treatment can have both individual and population-level impacts on health. For example, fast diagnosis and effective treatment of *Plasmodium falciparum* malaria can lead to population-level reductions in disease transmission (Landier et al., 2016). Conversely, delayed diagnosis and treatment of malaria can lead to increased severity of illness, worse clinical outcomes, cognitive impairment, and increased maternal and infant mortality (Landier et al., 2016). Similarly, the delayed diagnosis at the health facility of other diseases, for example, cholera cases has been associated with the onset of cholera outbreaks in various countries (Ratnayake et al., 2020).

Moreover, timely access to healthcare facilities is crucial for health conditions outside of infectious diseases as well. Snakebite envenoming, currently listed as a neglected tropical disease, impacts about 5.4 million people yearly, leading to approximately 138,000 deaths (*Snakebite*, 2022). Urgent medical attention is vital, particularly in rural areas. Vulnerable groups like workers, children, and those with restricted healthcare access face heightened risk (Appiah, 2012; Iliyasu et al., 2015). Prompt access to healthcare can mitigate these impacts and improve health outcomes. It is therefore key that geographic access is evaluated periodically, and that inequities are addressed, to ensure communities are within acceptable travel time and distance of healthcare providers.

Seasonal variation can significantly impact accessibility of essential medical services. For instance, Makanga et al. employed spatiotemporal modeling in Mozambique, revealing a significant drop in the number of women within two hours of life-saving care during both the dry and wet seasons (2017). Blandford et al. studied health facility access in Niger, observing a decrease in the proportion of the population within a one-hour walk to a health center during the wet season compared to the dry season (2012). Likewise, Ouma et al. found fluctuating proportions of the population within one-hour travel times for healthcare across different regions in Uganda during both dry and wet periods (2021). These studies underscore the importance of considering seasonal variations to comprehensively address healthcare accessibility challenges and design more effective strategies for ensuring timely access to maternal care and other health services throughout the year.

Periodic evaluation of healthcare access can be important for data-driven planning of the location of healthcare facilities (Moturi et al., 2022). Evaluating variations in geographic access is especially important because healthcare resources are often unequally distributed. Moreover, communities located more than two hours away from the nearest emergency healthcare facility face heightened risks during crises, aligning with the standards set by the World Health Organization and Lancet Commission for Global Surgery(Esquivel et al., 2016; Walther et al., 2020).

A range of studies across SSA have analysed accessibility to healthcare. The few studies that have evaluated the impact of precipitation and floods on access found that travel time increased and accessibility decreased during the rainy season in SSA (Blanford et al., 2012; Dotse-Gborgbortsi et al., 2022; Makanga et al., 2017). However, previous related work in Côte d’Ivoire has not accounted for weather seasonality.

In this study, we model and analyse geographic accessibility to public healthcare facilities in Côte d’Ivoire while accounting for seasonality, both during rainy and dry seasons. We quantify the proportion of the population that has poor access to timely healthcare given different seasonality. Finally, we investigate the spatial clustering of travel time at the health district level to determine the cold and hotspots of the population with poor access to healthcare.

## Methods

### Country context

Côte d’Ivoire, a lower middle-income country, located in West Africa shares its southwestern border with Liberia and its northwestern border with Guinea. To the north and northeast, it is adjacent to Mali and Burkina Faso, while to the east, it is bordered by Ghana. The country had a projected population of approximately 27.4 million people in 2019 (*World Bank Open Data*, 2022). The country exhibits diverse and heterogeneous human settlement patterns. The most densely populated areas include the economic capital-Abidjan, the central region surrounding Yamoussoukro, and the coastal areas along the Gulf of Guinea with more than 1,000 people per square kilometer. In contrast, the northern and western parts of the country have lower population densities with fewer than 100 people per km^2^ in the different districts. The population distribution has implications for healthcare infrastructure, demand for services, planning, and the local disease burden (Chen et al., 2023). As of 2021, 53% of the population (around 15 million people) lived in urban settings (UN-Habitat, 2023). Major urban centers include Abidjan, Bouaké, Daloa, San Pedro, and Yamoussoukro.

The provision of healthcare in Côte d’Ivoire involves a mix of governmental, non-governmental, and faith-based organizations, and private-for-profit managed health facilities (Cisse, 2010). The healthcare system follows a hierarchical structure with multiple tiers. At the community level, community health units provide basic health services. Primary care facilities, such as dispensaries, clinics, and health centers, form the next tier, providing more comprehensive healthcare services. County referral hospitals serve as the first and second referral points for specialized care within specific regions, while national referral hospitals offer tertiary care services. In Côte d’Ivoire, the 33 regional health departments serve as administrative units overseeing healthcare planning and policy implementation, while the 113 health districts manage healthcare service delivery at the local level, ensuring accessibility to medical services and public health programs within communities (President’s Malaria Initiative [PMI], 2023). Furthermore, the 510 subprefectures are responsible for governance and administration within specific geographical areas. They serve as intermediaries between the central government and local communities, overseeing functions such as civil registration, law enforcement, and public services.

The correlation between travel times in Côte d’Ivoire and weather conditions is closely tied to healthcare-seeking behavior. Road types play a pivotal role, with major roads—less vulnerable to rain—contrasting significantly with smaller roads, walking, and bicycling speeds, which experience notable delays. Furthermore, the transportation mode to healthcare facilities is associated with the resources available to the individuals. In 2015, The Survey on Household Living Standards in Côte d’Ivoire 2015 (ENV 2015), underscored the prevalence of walking for accessing primary healthcare (48.8%), while public transportation is favored for reaching general hospitals (24.9%), demonstrating socio-economic disparities (INS, 2015). Further research into healthcare utilization in Cote d’Ivoire revealed that proximity to healthcare providers influences health-seeking behavior. This impact is especially evident in regions connected by smaller roads, where rainy seasons result in substantial increases in travel times. To capture this trend, our study adopted a reduced travel speed approach, halving all travel speeds during wet periods.

The country has witnessed significant governance and healthcare transformations since the early 2000s (Gaber & Patel, 2013). The quality of healthcare and the efficiency of the health system are still recovering from years of conflict. Political and constitutional reforms have led to decentralization, establishing regions and autonomous districts. As a result, healthcare management responsibilities, including facilities and service delivery, now lie with regional and district authorities. The Ministry of Health and Public Hygiene maintains oversight of national healthcare policies, regulations, and management of national referral hospitals. These reforms align with the Côte d’Ivoire 2016 Constitution, which recognizes the right to the highest attainable standard of health (*Côte d’Ivoire 2016 Constitution - Constitute*, 2016). Health policy priorities focus on improving access to primary healthcare, ensuring high-quality services, strengthening infrastructure, enhancing workforce capacity, and promoting equitable healthcare access nationwide.

Malaria, HIV/AIDS, and tuberculosis are the country’s leading causes of morbidity and mortality, and non-communicable diseases such as cardiovascular diseases and cancer are becoming more prevalent (Cisse, 2010; Ilesanmi et al., 2021). The National Health Development Scheme (PNDS) is in charge of the delivery of healthcare services, but the public health sector is underfunded.

### Overall geospatial analysis framework

The overall analysis framework is provided in Figure 2. Health facilities, population distribution, and factors that affect travel were assembled first. A travel time scenario for each season was based on literature review and used together with the assembled data to estimate both the travel time and the proportion of the population within specific travel time threshold to the nearest public healthcare facility. (Fig. 2)

**Figure 2.**
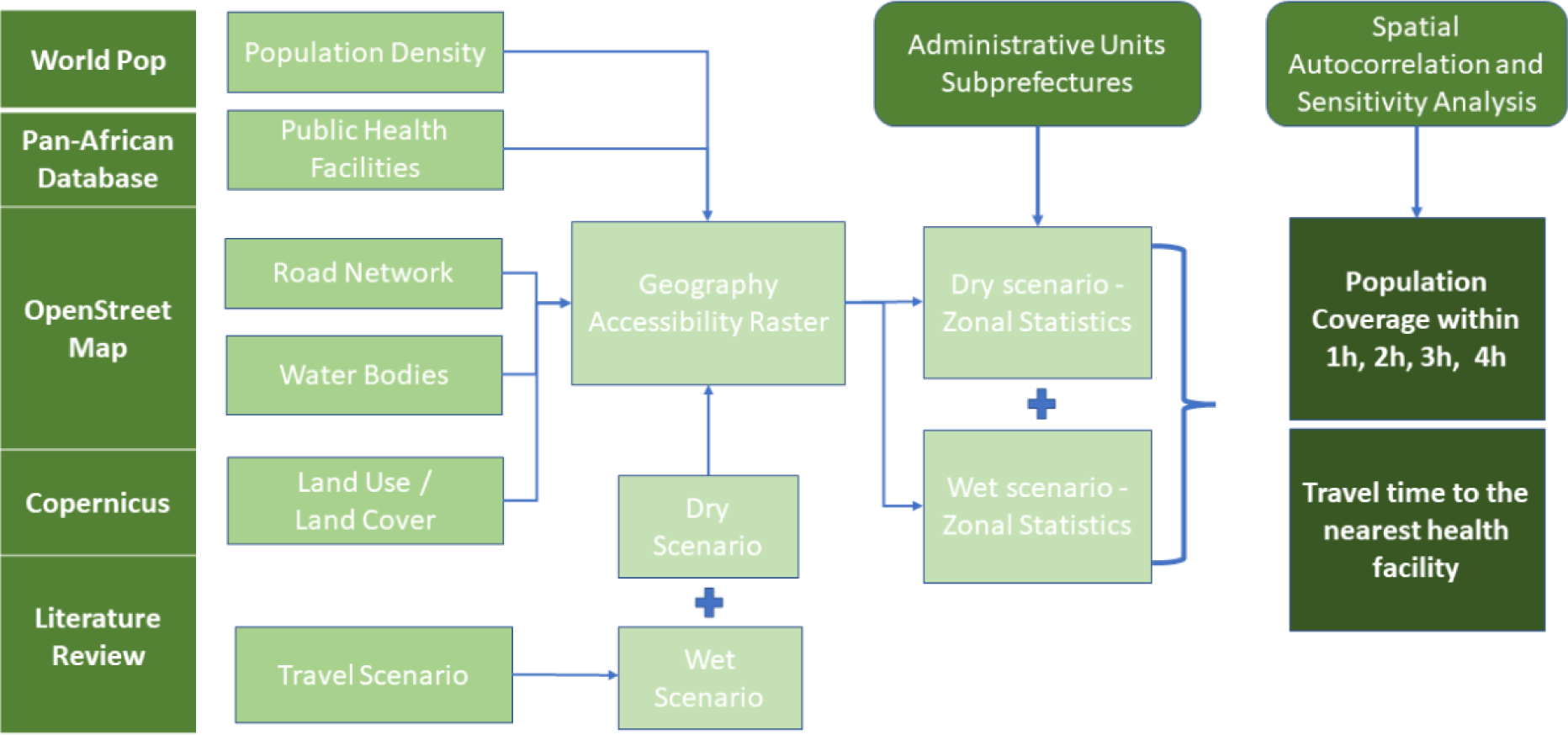
Overview of the geospatial modelling framework to compute travel time to the nearest public health facility in both wet and dry seasons in Cote d’Ivoire.

### Data

#### 1. Geocoded database of public health facilities

Previous work generated the first spatial census of public health facilities in SSA(Maina et al., 2019). Based on this list, within Côte d’Ivoire, we extracted public health facilities (government, faith-based, and non-governmental facilities) and their corresponding Global Positioning System (GPS) coordinates. We randomly sampled 80% of the health facilities and validated their locations via Google Earth. Based on the extracted list, Côte d’Ivoire has 1,660 health centers, 96 national hospitals, and 4 teaching hospitals (3 of which are located in the Lagunes region in south Cote d’Ivoire) (Figure 3. A.). All the country’s regions have at least one regional hospital, except for the Lagunes and Bandaman Valley regions. Instead, there are 11 national hospitals in the Lagunes region and 5 in Bandaman Valley. Baffing and Denguélé regions (north) have no national hospitals.

**Figure 3.**
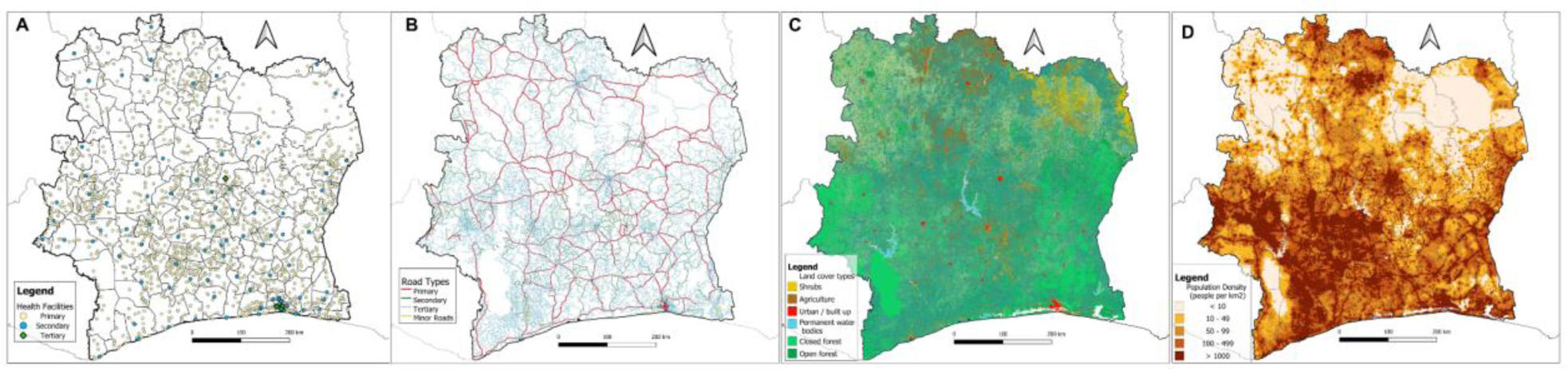
**A.** Locations of Public Health Facilities disaggregated by level derived from the pan-African database of health facilities. **B.** Road Networks derived from OpenStreetMap. **C.** Land Cover/ Land Use type derived from Copernicus. **D**. and the spatial distribution of population in Cote d’Ivoire. Regions in light colors are inhabited national parks in Côte d’Ivoire [Comoé National Park in the northeast; Parc National du Banco on the coastal; Taï National Park South-west]

#### 2. Road Network

When seeking healthcare, patients tend to use roads for walking or driving more often than other types of land areas. Therefore, we assembled a comprehensive database of road networks with the entire country. These data were downloaded from OpenStreetMap (OSM) where information on walking, bicycling, and motorized travel, including different types of roads were included. The roads were recategorized as primary, secondary, and tertiary roads. Those classified as primary were high-volume roads that mainly connect international borders. Secondary roads were those that fed into primary roads or connected major towns in different regions of a country. Tertiary roads connected secondary roads while connecting smaller towns or market centers. (Figure 3.B.)

#### 3. Land Use / Land Cover

Where no road network existed, land cover/ land use (LULC) was used to designate landscapes that people need to traverse when accessing healthcare. The LULC layer was derived from Copernicus (Buchhorn et al., 2020) and included shrubs, grassland, herbaceous vegetation, cities, and water bodies as land types. (Table 2) The water bodies from the LULC layer were augmented with river (line data) data derived from OSM. We assumed that water bodies act as barriers, except where a road intersects the water body. (Figure 3.C.)

**Table 2.** Summary of Travel Times on different road surfaces and on different land cover types in Côte d’Ivoire during wet and dry seasons.

|  | Ouma et al., 2021 | Blandford et al., 2012 | Makanga et al., 2017 | Current study |
| --- | --- | --- | --- | --- |
| **Dry [Wet]** |  |  |  |  |
| Primary (km/h) | 50 [40] | 30 [20] | 120 [96] | 100 [50] |
| Secondary (km/h) | 30 [24] | 17 [8] | 80 [64] | 50 [25] |
| Tertiary (km/h) | 30 [24] | 17 [8] | 40 [32] | 30 [15] |
| Walking Pathways | 10 [8] | 2- 5 [1.7 - 4] | 3-5 [2-4] | 10 [5] |
| **Land cover type** |  |  |  |  |
| **Open or sparse vegetation**<br>shrubs, open forest, closed forest. | 2-5 [1.6 - 4] | 3 [2] | n/a | 2 - 4 [1 - 2] |
| **Deciduous Shrubland /woodland**<br>Agriculture | 4-5 [3.2 - 4] | 1.5 [1] | n/a | 2 [1] |
| **Water Bodies** | 0.01 [0.01] | 0 [0] | Impassable | 0 [0] |
| **Urban/ Built up** | 5 [4] | 4 [3] | n/a | 5 [3] |

#### 4. Population distribution

We obtained estimates of the population density for the entire nation at a spatial resolution of 1km^2^ from the WorldPop database for 2020 (Stevens et al., 2015). To construct the high-resolution population gridded surface, WorldPop uses the latest census data and disaggregates based on land use and land cover grids. The various land use types and cover categories were assigned weights based on the likelihood of having a human population. A random forest method is employed to perform the breakdown, accounting for differences between rural and urban areas, and the UN’s population growth projections for urban and rural regions of Côte d’Ivoire. (Figure 3.D.)

### Health care seeking during dry and wet seasons

We modelled travel time to public health facilities based on transport scenarios that included motorized transport, bicycling, and walking based on literature (Ouma et al., 2018; Walther et al., 2020) and information from OSM attribute data (Table 2.). Different speed limits were assigned to different land types and roads for two overarching models: one model considering a travel scenario during dry season (assuming that all individuals can travel at relatively high speeds uninterrupted) (Table 2.) and a second model considering a reduced travel speed scenario (with all speeds from the first model reduced by half). These assumptions are based on studies that have considered seasonality while computing spatial accessibility in SSA previously (Delamater et al., 2012), Ouma et al. (2021), Blandford et al. (2012), and Makanga et al. (2017)). (Table 2)

### Travel time computation during dry and wet seasons

Travel time estimation was done using AccessMod version 5.8. AccessMod is a WHO open-source standalone software that uses a cost-distance algorithm to model how physically accessible existing health services are to the target population. First, the land cover, rivers, and road layers were merged to create a friction raster surface at 100m spatial resolution. This friction surface was used in combination with the point locations of healthcare facilities, to generate a raster layer of anisotropic travel times to the nearest healthcare facility for the entire nation using the *Accessibility Analysis function*. The function computed cumulative travel time from each 100m grid to its nearest facility. The analysis was confined within the national borders, assuming populations do not cross-national borders to use health facilities in neighboring countries. We ran two models, one during the wet season and the other during the dry season based on speeds in Table 2. For policy purposes, we aggregated the high-resolution estimates at the district level.

### Geographic coverage

To determine the proportion of the population within the recommended travel threshold at the health districts level we used the “Zonal Statistics” function in AccessMod along with the WorldPop population density raster layer. The function aggregated the population that has access to healthcare at different time increments (1h, 2h, 3h, 4h) and mapped using QGIS (version 10.5).

The choice of 1-hour and 2-hour time increments for routine care is based on recommendations from the Lancet Commission on Global Surgery, which defines geographical accessibility in terms of the proportion of the population that can reach a facility with essential surgical care within 2 hours (Pérez-Soto et al., 2023). We included 3-hour and 4-hour time to identify individuals who face significantly increased challenges in accessing healthcare (extremely marginalized). By considering a range of time increments, from 1 to 4 hours, we provide a comprehensive assessment of healthcare accessibility, taking into account both emergency and routine care needs.

### Spatial autocorrelation

We used spatial autocorrelation statistics to assess the existence, and locations, of spatial clustering in poor geographic accessibility to healthcare facilities. The clustering statistics were run on the estimated percentage of the population that is more than: four hours; three hours; two hours; and one hour of travel time to the nearest facility for every health districts and subprefectures. We first used the global Moran’s I statistic to assess spatial autocorrelation across the entire nation, while the Anselin’s Local Indicators of Spatial Autocorrelation (LISA) was used to quantify and map hotspots and coldspots within the country (Anselin, 1995). The Moran’s I statistic indicates the degree of spatial clustering whereas the simulated p-value indicates statistical significance. The values range from roughly −1 to 1, with 1 indicating complete spatial clustering (i.e., all areas with high values are neighboring other areas with high values) and −1 indicates complete spatial dispersion (with high-value areas always neighboring low-value areas). The LISA statistics are a local version of the Moran’s I, with a separate clustering or dispersal statistic and p-value estimated for each administrative unit, and the results most commonly interpreted by mapping and visualization. We used a Bonferroni correction to account for multiple testing (Armstrong, 2014). The Bonferroni correction addresses the issue of multiple hypothesis testing, reducing the risk of false positives by adjusting the significance level for the number of tests conducted.

## Results

### Travel time

On average, at the national level, the travel time to the nearest public health facility was 1.8 hours in the dry season and 3.4 in the wet season. Notably, travel times showed considerable heterogeneity across the districts particularly along the coast, ranging from 23 minutes to 6 hours in the dry season and 38 minutes to 10 hours during the wet season (Figure 7. C and D). 59 districts had a mean travel time of more than 2 hours in the dry season (Figure 7.C), this increased to 85 districts during the wet seasons (Figure 7.C). These marginalised districts are located in north and west areas in the dry areas which shift to most of the country in the wet season. (Figure 7. A and B).

**Figure 7.**
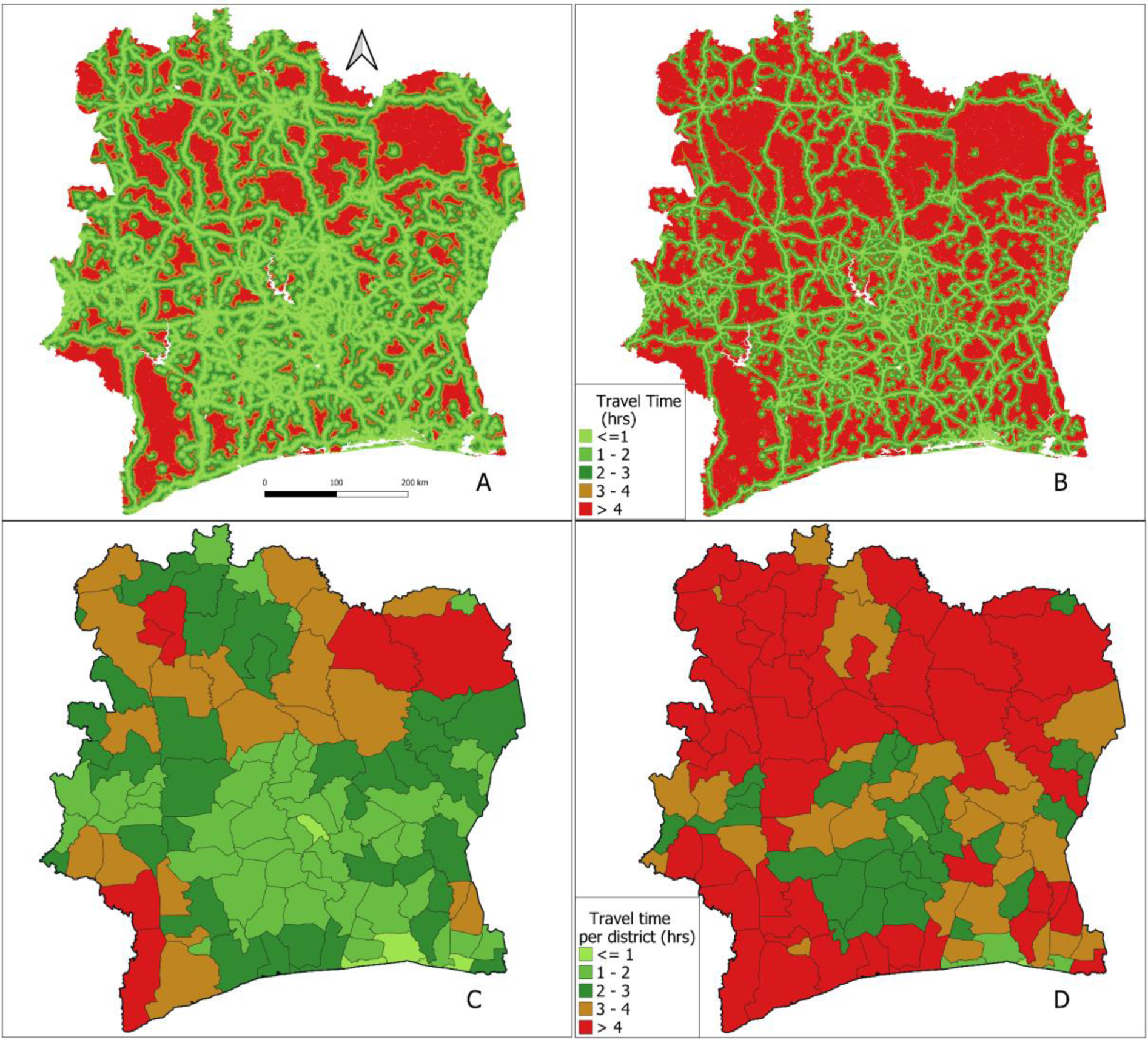
Travel time to the nearest health facility during the dry (A and C) and wet seasons (B and D) at 100m spatial resolution (A and B) and at mean travel time per health district (C and D)

### Geographic Coverage

The results demonstrate varying levels of mean estimated percentage of the total population within 1, 2, 3 and 4 hours travel time to the nearest healthcare facility at the national level during dry and wet seasons. In the dry scenario, approximately 64% of the population can reach the nearest healthcare facility within 1hour, which increases to 80% within 2 hours, 88% within 3 hours, and 93% within 4 hours. Conversely, during the wet season, the accessibility slightly decreases, with 53% of the population within 1h, 66% within 2h, 75% within 3h, and 83% within 4h. These results highlight the seasonal variability in healthcare accessibility, with generally higher accessibility during dry periods compared to wet seasons.

Significant heterogeneity in healthcare access was evident across the country and seasons. For example, using the 2-hour threshold in dry settings, population coverage ranged from 7 to 98%, averaging 73% across all health districts. In wet settings (still looking at the percentage of the population within 2 hours to the nearest facility), coverage ranged from 2 to 97%, averaging 55%. Throughout all timeframes, access was consistently better in dry weather than in wet weather (Figure 8. A & B.). For instance, two health districts: Jacqueville and Abidjan both located in the south, had in Jacqueville (south) 9% versus 36% (dry versus wet) of the population were more than 2 hours away from a health facility, while in Abidjan (south) it was 8% versus 10% similarly in the south. Similarly, in the north, higher percentages of the population were more than 2 hours away from healthcare facilities. Odienne (northwest) had 37% versus 54% (dry versus wet) of their population more than 2 hours away from the nearest healthcare facility. Madinani (northwest) had 55% versus 72% (dry versus wet) of their population more than 2 hours away from the nearest healthcare facility (Figure 8. A & B.).

**Figure 8.**
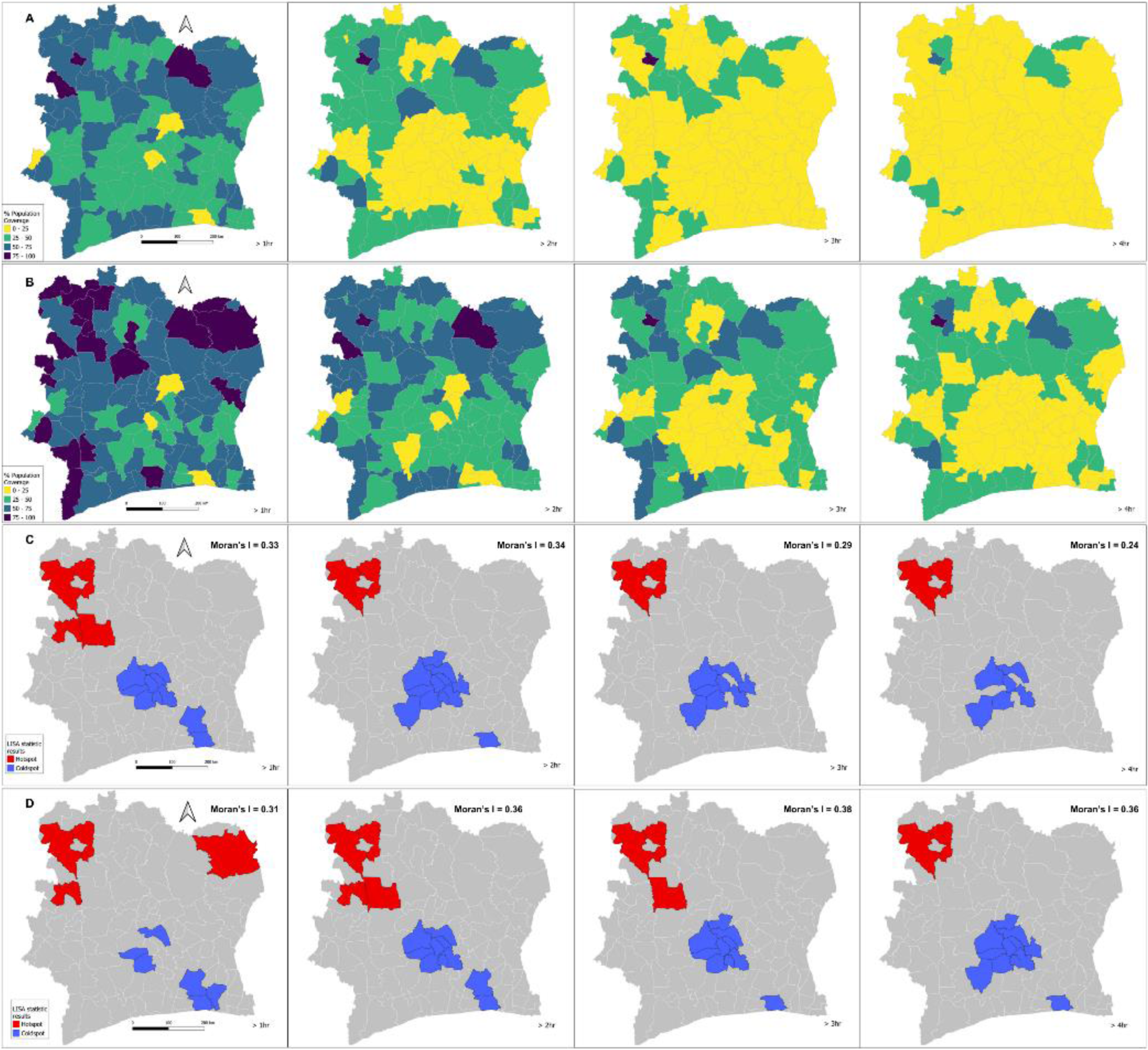
Percent of Population Coverage with poor access to healthcare per health districts at 1, 2, 3, 4 hours time increment in dry **A.** and wet **B.** settings. Spatial clustering hotspots (blue) and cold spots (red) Health Coverage at 1, 2, 3, and 4 hours time increment in dry settings **C.** and wet season **D.**

### Clustering

The spatial autocorrelation analysis identified distinct hotspots and coldspots of healthcare facility access. In this analysis, hotspots indicate clustering of administrative units with poor access to healthcare facilities whereas coldspots indicate clusters of administrative units with good access to healthcare facilities. In dry settings, across the 1 to 4-hour travel time intervals, persistent hotspots were evident in the northwestern region, while coldspots were observed in the central and southern areas. Hotspots are regions where the population has limited access to healthcare facilities, indicating a concentration of areas with relatively higher travel times to reach medical services. While coldspots are areas where access to healthcare facilities is better, with the population in these regions having shorter travel times to reach medical services. In wet settings, a similar pattern emerges, with consistent hotspots persisting in the northwestern region within the 1 to 4-hour intervals. Coldspot areas in the central region expand over time, along with their prevalence in the south. (Figure 8. C & B.)

For both scenarios, Moran’s *I* values were positive and statistically significant, indicating spatial clustering in the percentage of the population within different time intervals of the nearest healthcare facility. Across all time intervals exceeding 1, 2, 3, and 4 hours, both scenarios exhibit statistically significant spatial clustering, with p-values equal to 0.001. Notably, the “Wet Scenario” consistently reports higher Global Moran’s *I* values compared to the “Dry Scenario.” This data underscores the presence of spatial clustering in accessibility to healthcare facilities, with the “Wet Scenario” consistently reflecting stronger clustering patterns in comparison to the “Dry Scenario.”

## Discussion

This study modelled geographic accessibility to public healthcare facilities across Côte d’Ivoire, factoring in reduced travel capacity during the rainy season compared to the dry season (Blanford et al., 2012; Makanga et al., 2017; Ouma et al., 2018). Significant disparities in accessibility were observed across seasons and regions, with concentrated poor accessibility in the northern regions and better accessibility in the southern regions, particularly along the coast. These disparities may be associated with limited transport networks during wet seasons, inadequate infrastructure in remote areas, limiting transportation networks and traffic congestion in urban areas in Côte d’Ivoire (Agbenyo et al., 2017; Carrasco-Escobar et al., 2020; Weiss et al., 2018).

We demonstrated that the travel times to the nearest health facilities increase twice more during the wet season from 1.8h to 3.4h. Notably, during the dry season, approximately half of the districts (59 out of 113) reported travel times exceeding 2h, these number changed during the wet season and two-thirds of the districts (85 out of 113) reported travel times exceeding 2h. Moreover, in dry conditions, 64% of the population could access a health facility within 1h, extending to 93% within 4h. In contrast, during the wet season, coverage dropped to 53% within 1 hour and 83% within 4h.

Our results could be driven by the various geographical features, such as the elevated terrains and dense forests in the north-western regions. For example, the western regions of Côte d’Ivoire, including Moyen-Cavally and Montagnes, are characterized by dense forests and hilly terrain. In the north-western regions, the road infrastructure has historically posed difficulties, with remote villages accessible only via rough, unpaved roads, further extending travel times. Moreover, densely populated areas typically boast better infrastructure and services, resulting in shorter travel times. In contrast, sparsely populated regions, such as Tchologo and Bagoué, face significant challenges in establishing and maintaining healthcare facilities and transportation networks due to lower population densities compared to urbanized hubs like Abidjan (The World Bank, 2022).

The accessibility to healthcare facility is further influenced by the distinction between rural and urban areas. Conversely, urban areas contend with improved road networks, but it becomes overwhelmed during the rainy seasons, resulting in extended travel times. This stark divide highlights the intricate interplay between population density, urbanization, and geographic accessibility in shaping healthcare access in Côte d’Ivoire. Both rural and urban areas face distinct challenges related to geographic accessibility, as healthcare resources tend to be concentrated in urban centers, leaving rural regions with limited access to quality medical care; consequently, in rural areas, healthcare facilities are frequently situated far from the population, and during the rainy seasons, unpaved roads become difficult or impossible to traverse (Blanford et al., 2012; Dotse-Gborgbortsi et al., 2022). On the other hand, urban areas are affected by high traffic congestion resulting in longer travel times, informal settlements with poor communities, and poor road infrastructure that are also affected during the rainy season (Poku-Boansi, 2021).

As of now, Côte d’Ivoire does not have universal healthcare coverage. Individuals must often pay for out-of-pocket expenditures, which turns out to cost more in private clinics compared to public health facilities. In 2018, the utilization rate of public sector health services was estimated to be 47% (Duran et al., 2020). The utilization rate of public health facilities is low and turns out to be lower in northern subprefectures where resources (monetary, human, medicinal) are lacking. Building upon prior research on travel time assessments, this study introduces a novel dimension by examining heterogeneities in accessibility across the nation. Such insights are crucial in a country grappling with elevated maternal and infant mortality rates, largely attributable to limited healthcare access. The study also underscores the multifaceted nature of accessibility disparities, influenced by geographic, ecological, historical, and individual factors. Efforts by public health programs and partnerships are laudable steps towards improving accessibility, particularly in the realm of malaria prevention and postpartum family planning.

During the rainy seasons in Cote d’Ivoire, regions along the coast and in the southern part of the country are particularly prone to flooding. For instance, in Abidjan and neighbouring regions, 11,478 people were affected by heavy flooding in this area in 2022. (International Federation of Red Cross and Red Crescent Societies (IFRC), 2022). The risk of waterborne diseases such as malaria, cholera, and yellow fever is high. And the difficulty of accessibility increases because bridges may be destroyed, and roads are impassable. The interplay between accessibility and waterborne disease outbreaks after heavy rainfall serves as a stark reminder of climate change’s profound impact on public health. The escalation in frequency and intensity of extreme water-related events, driven by global climate shifts, significantly heightens the vulnerability to waterborne outbreaks. Notably, pathogens like *Vibrio* spp. and *Leptospira* spp. emerge as prominent concerns in these heightened risk scenarios. This underscores the critical need for proactive and equitable measures in bolstering geographic accessibility, especially in regions susceptible to environmental challenges. Such strategic enhancements in accessibility hold the potential to fortify public health resilience in the face of evolving climatic conditions (CANN et al., 2013; Shih et al., 2021).

This work demonstrates how the presence of natural obstacles, such as dense forests, hilly terrain, and bodies of water, poses significant challenges for transportation. To address these disparities, policy actions should prioritize equitable resource allocation, deploy mobile clinics with telemedicine integration, establish emergency transport networks, enhance transportation infrastructure, and foster cross-sector collaboration, especially in rural areas. These measures, combined with data-driven decisions, public-private partnerships, and healthcare infrastructure upgrades, will work synergistically to bridge accessibility gaps and foster inclusivity. An example of a program that improves geographic access to healthcare in Cote d’Ivoire is The Inclusive Connectivity and Rural Instracture Project. The project, funded in 2023, aims to improve the road infrastructure in the Northern region to reduce transport times along road sections and improve access by ambulances to rural health centers for the transfer of patients to referral hospitals. (African Development Bank (AFDB)Group, 2016; Asian Infrastructure Investment Bank (AIIB), 2023).

## Limitations

Our study has several limitations. Our model focuses on geographic accessibility proximity (Guagliardo, 2004; Kahabuka et al., 2011). This approach overlooks other potentially important barriers to accessing public health facilities. Physical proximity to a healthcare facility does not guarantee access to care, especially if the cost of care is prohibitively high for the patient. Our results serve as indicators of access to primary services, and do not offer a universally applicable assessment of healthcare access in all circumstances. The second limitation is the estimation of travel speed and travel time (Deng & Bennett, 2023). We borrowed the values from prior research on access to care, we note that there is a lack of empirical evidence demonstrating their applicability to our specific study setting. We want to emphasize that our study considered variability due to climatic factors but did not incorporate precipitation data. While these additional analyses fall outside the scope of this paper, it is important to acknowledge their potential importance in advancing our understanding of healthcare service access across diverse domains.

## Conclusion

The study illuminates the disparities in geographic accessibility to public healthcare facilities across Côte d’Ivoire, shedding light on the pronounced challenges faced, particularly in the Northern regions, exacerbated during the rainy season. These findings emphasize the critical need for targeted interventions to bridge these regional gaps, especially in the context of flooding events, which further exacerbate access limitations. Geographic accessibility is important for timely diagnosis, reducing morbidity, and improving maternal and child health. As such, stakeholders and policymakers must prioritize strategies that not only address geographical barriers but also consider the impact of natural disasters, such as floods, on healthcare accessibility. This integrated approach will be pivotal in building a resilient healthcare system capable of withstanding the challenges posed by both geographical and environmental factors.

## Supporting information

Supplementary informaiton

## Data Availability

All data produced in the present study are available upon reasonable request to the authors

## Notes

### Competing Interest Statement

The authors have declared no competing interest.

### Funding Statement

This study did not receive any funding

