## Supplementary informaiton for "Geographic accessibility to public healthcare facilities and spatial clustering during the wet and dry seasons in Côte d’Ivoire"

**Supplementary information**

**Table 1.** Categories and levels of Public Health Facilities in Côte d'Ivoire derived from the pan-African database of health facilities.

| **Type of Health Facility** | **Number of health facilities** | **Hierarchy** |
| --- | --- | --- |
| Rural Health Center | 1331 | **First Contact (Primary)** |
| Urban Health Center | 329 |  |
| General Hospital | 77 | **First Referral (Secondary)** |
| Regional Hospital | 19 |  |
| University Hospital | 04 | **Second Referral (Tertiary)** |

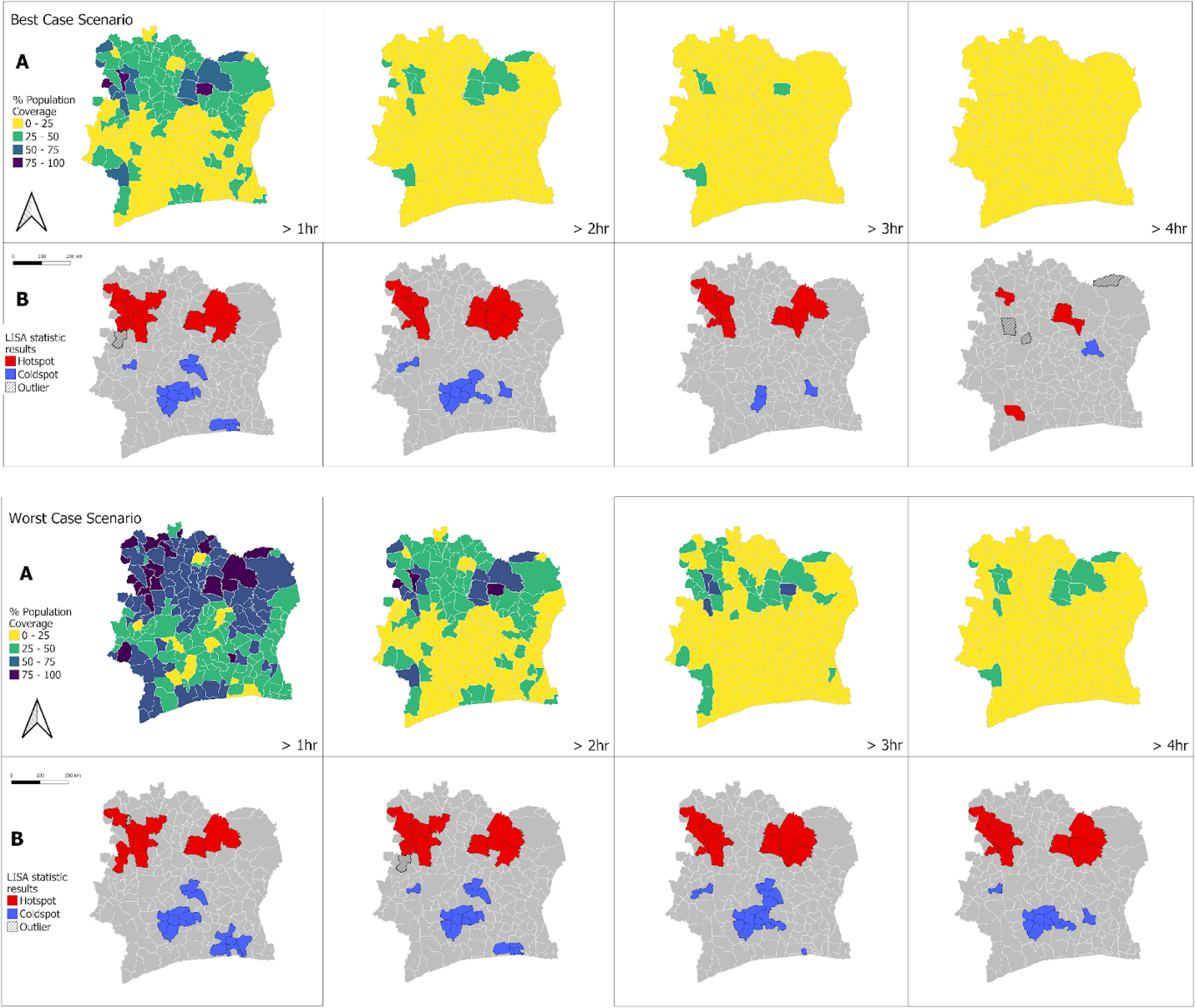

**Figure 9.** Percent of Population Coverage and Spatial Clustering in Cote d’Ivoire per subprefectures. A. Population Coverage with poor access to healthcare at 1, 2, 3, 4 hours time increment. B. Spatial clustering Good (blue) and Bad (red) Health Coverage at 1, 2, 3, 4 hours time increment.

At the subprefecture level we found extreme heterogeneity in the distribution of access to healthcare facilities across the landscape - ranging from 27% of the population that is greater than 4 hours away from a facility to less than 1%. A cluster of regions with relatively poor access to healthcare facilities exists across the northern area of the nation. Northern regions such as Worodougou and Bafing Denguele had high percentages of the population being farther than 4 hours from the nearest healthcare facility ranging from 20.07% to 26.59%. Meanwhile, a cluster of areas with better coverage of healthcare facilities exists in the central coastal areas (around Abidjan) and extends into the central part of the nation where only 1% of the population is further than 4 hours from the nearest healthcare facility. Additionally, we found that regardless of the time travel and the setting (dry vs wet) the number of individuals that did not have access to any public health facilities in the north region was highest. (Figure 9. A)
